# Enhancing Clinical Reasoning in Medical Vision-Language Model through Structured Prompts

**DOI:** 10.1101/2025.07.31.25331901

**Authors:** Kavya Dasaramoole Prakash, Kiseong Kim, Youngmahn Han

## Abstract

Medical Vision-Language Models (MVLMs) are emerging as powerful tools for tasks such as Visual Question Answering (VQA); however, they often struggle with hallucination and limited reasoning transparency, particularly in complex diagnostic scenarios. In this work, we enhance the MedVLM-R1 framework by fine-tuning it using clinically informed prompt structures tailored specifically for radiology-based reasoning. Without altering the original model architecture or training strategy, we redesign the system prompts and question templates to guide the model through structured, modality-aware, and step-by-step diagnostic reasoning. Fine-tuning is performed using MRI-based question-answer (QA) pairs, and evaluations are conducted across three diagnostic imaging: MRI, CT, and X-ray to assess both in-domain and out-of-domain generalization. Our approach improves reasoning transparency and accuracy, achieving 96.00% on MRI, 72.67% on CT, and 75.2% on X-ray. Compared to the original MedVLM-R1, our method closes the gap in MRI accuracy while significantly enhancing generalization performance on CT and X-ray modalities. These results demonstrate that clinically grounded prompting effectively improves both reasoning fidelity and robustness across imaging modalities. The code is available at our GitHub repository:https://github.com/aidanbio/AIdanMed

## 1 Introduction

In recent years, language modeling has made remarkable progress. Large Language Models (LLMs) have shown the ability to handle an extensive number of tasks, driving their growing popularity. Originally designed for text-only inputs, these models are now being extended to process visual information as well. Integrating vision with language opens the door to numerous applications that are central to the ongoing AI-driven technological transformation [1]. Vision-Language Models (VLMs) mark a significant breakthrough in artificial intelligence, facilitating multimodal understanding and reasoning by bridging textual and visual data. MVLMs mark a significant breakthrough in AI-powered healthcare, with the potential to improve the analysis of medical images, automate report generation, and support clinical decision-making [2]. Recent initiatives aim to enhance the capabilities of VLMs by incorporating explicit reasoning methods that prioritize interpretability and step-by-step explanation. Chain-of-Thought (CoT) prompting is a popular strategy for guiding models to generate intermediate reasoning steps prior to obtaining a final solution [3].

Although CoT prompting increases reasoning transparency in VLMs, training procedures are also important in aligning model behavior with clinical reasoning. Conventional fine-tuning techniques, such as Supervised Fine-Tuning (SFT) [4, 5], frequently rely on static labeled datasets, which may not adequately capture the complex reasoning or diverse interpretations needed in clinical situations. To address this limitation, Reinforcement Learning (RL)-based techniques [6] have gained popularity, allowing models to iteratively adjust their outputs based on feedback signals that emphasize correctness, coherence, and clinical relevance. Group Relative Policy Optimization (GRPO) [7] is a promising RL approach for increasing reasoning accuracy and answer quality in multimodal challenges. Unlike conventional RL or SFT, GRPO employs a group-based comparison reward structure, allowing the model to optimize not just for the single best response but also for the relative quality of sampled candidate solutions. This helps the model generalize more effectively across clinical circumstances and varying levels of reasoning depth.

One of the most recent developments is MedVLM-R1 [8], a powerful vision-language model designed for medical use. MedVLM-R1, built on a strong multimodal foundation, combines instruction-tuned language models with visual encoders and uses reinforcement learning strategies like GRPO to improve reasoning. It has shown remarkable performance on clinical tasks such as generating radiology reports and VQA, especially when using group-based reward optimization techniques with CoT prompting.

Nevertheless, MedVLM-R1 has several drawbacks despite its advantages, most notably its dependence on generic system prompts that are neither task-specific nor clinically grounded. Especially in complex imaging scenarios involving multiple modalities, this often results in outputs that are semantically fluent but clinically imprecise or underexplained. Previous research has also documented cases of unsupported conclusions or hallucinated reasoning [9], in which the model produces explanations that seem convincing but lack clinical validity. Better alignment of model inputs and training signals with real-world diagnostic expectations remains critical, as medical decision-making necessitates precise, fine-grained, and context-aware reasoning.

### 1.1 Previous Work

a. **General Vision-Language Models**: The foundation for multimodal comprehension was established by early developments in VLMs like BLIP [10], Flamingo [11], and LLaVA [12], which aligned visual and textual modalities through extensive pretraining on interleaved image-text datasets. These models frequently achieved competitive performance in zero-shot and few-shot conditions, exhibiting strong generalization performance across multiple downstream tasks, including visual question answering, captioning, and instruction-based tasks. However, although general-purpose VLMs perform well on open-domain tasks, they perform poorly in domain-specific contexts such as healthcare, where medical expertise and deeper reasoning are crucial. The complex visual patterns and textual semantics present in healthcare settings are frequently missed due to their generic training material and prompts.
b. **Medical Vision-Language Models**: To address the domain gap, several medical-specific VLMs have been developed. Models such as BioViL [13] and MedCLIP [14] used contrastive learning to match radiological images with corresponding reports, enhancing visual representations in clinical settings. With more varied training signals, BioMedCLIP [15] expanded this method to larger biomedical corpora, improving cross-modal comprehension. Additional developments include instruction-tuned models that use CoT reasoning to provide clinically appropriate outputs, including LLaVA-Med [16], Med-PaLM 2 [17], and Med-R1 [18]. These models demonstrate the value of combining structured prompting with medical knowledge by achieving strong results on benchmarks such as VQA-RAD and PathVQA [19] [20]. Despite these strengths, the majority of medical VLMs continue to use general system prompts and static evaluation, which may restrict their capacity to adapt to more complex, ambiguous clinical situations. Although medically credible, their results frequently lack contextual harmony with real-world decision-making processes and deeper reasoning.
c. **Reasoning with Chain-of-Thought and Reinforcement Learning**: To overcome these constraints, structured reasoning techniques like CoT prompting [3] have been developed. They encourage models to generate intermediate reasoning stages, thereby enhancing performance on challenging inference tasks. Approaches based on Reinforcement Learning (RL) have been proposed to overcome static tuning and fixed datasets. Techniques such as Reinforcement Learning with Human Feedback (RLHF) [21] and Direct Preference Optimization (DPO) [22] refine behavior in a task-aligned approach by optimizing models based on human feedback. GRPO introduces group-based reward comparison, enabling the model to learn from relative quality rather than absolute correctness. This approach has shown improvements in reasoning depth, output diversity, and clinical relevance. However, even with GRPO, performance depends heavily on the quality and specificity of prompts, especially in multimodal clinical scenarios. Generic instructions often fail to adequately direct the model toward reasoning paths that are task-specific or clinically appropriate.
d. **Prompt Design and Instruction Tuning in Clinical Tasks**: To mitigate these issues, recent work has emphasized the role of prompt engineering in guiding medical VLMs [23] [24]. Studies have shown that well-designed, clinically informed prompts improve interpretability, accuracy, and conformity to domain-specific expectations. Instruction tuning with structured prompts allows models to better simulate expert reasoning, enhancing their performance in diagnostic tasks, report generation, and VQA. In the following section, we build on these insights by enhancing MedVLM-R1 through domain-specific fine-tuning and radiology-grounded prompt design. We further evaluate the model’s reasoning consistency and generalization across modalities, demonstrating how targeted prompting can help overcome the limitations of generic instruction formats in producing accurate and interpretable outputs across diverse clinical scenarios.

### 1.2 Problem Statement

Previous research has also documented cases of unsupported conclusions or hallucinated reasoning [9], in which the model produces explanations that seem convincing but lack clinical validity. Improved alignment of model inputs and training signals with real-world diagnostic expectations remains critical, as medical decision-making necessitates precise, fine-grained, and context-aware reasoning.

To address these gaps, this work presents the following contributions:

i. **Fine-tuning of MedVLM-R1** using revised, clinically grounded prompts on existing domain-specific medical imaging datasets, enhancing diagnostic reasoning and improving performance across imaging modalities.
ii. **Clinically informed prompt design** that incorporates structured reasoning templates and modality-aware system instructions, designed to reduce hallucinated reasoning and improve alignment with expert clinical expectations.
iii. **Evaluation across X-ray, CT, and MRI modalities**, highlighting the effectiveness and generalizability of the proposed prompt modifications in producing accurate and interpretable outputs across diverse clinical scenarios.

## 2 Methodology

We build upon MedVLM-R1, a vision-language model developed for medical visual question answering, which integrates a Qwen2-VL-2B backbone [25]. To encourage robust and interpretable responses, we adopt GRPO, originally proposed in [7] and later employed in the medical domain by MedVLM-R1 [8]. GRPO [7] is an extension of Proximal Policy Optimization (PPO) [6] that replaces the traditional value function with a group-relative advantage computed from sampled outputs. This approach helps align model behavior with task-specific preferences while avoiding the instability of learned critics.

*G* candidate answers at every training stage 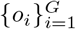 is sampled from the previous policy 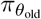 and evaluated using a corresponding reward function *r*_*i*_. The group-normalized advantage *A*_*i*_ is computed as:

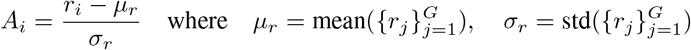

This encourages the model to prefer responses that are relatively better than others in the same batch, regardless of absolute reward scale.

The model parameters are then updated by maximizing the GRPO objective:

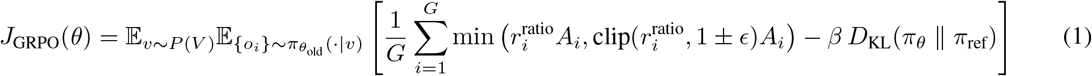

where 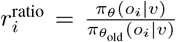 is the importance sampling ratio and *D*_KL_(*π*_*θ*_ ∥ *π*_ref_) is a KL-divergence penalty[26] that regularizes deviation from a reference policy *π*_ref_. The clipping threshold and KL regularization strength are controlled by the hyperparameters *ϵ* and *β*, respectively. This objective ensures preference alignment, robustness, and training stability across different modalities.

The reward function [27] used during training consists of two components:

1. **Format Reward**: This reward drives the model to adhere to the expected output format. Specifically, the model is required to enclose the reasoning in `<think>`…`</think>` tags and the final answer in `<answer>`…`</answer>` tags. If each of the four tags appears precisely once and no additional text appears beyond these tags, a reward of 1 is given. Any deviation from this, such as missing or duplicated tags or the presence of text beyond the tags, results in a reward of 0.
2. **Accuracy Reward**: Once the format is validated, the final answer within the `<answer>` tag is checked against the ground-truth label. If the model correctly selects the letter corresponding to the ground-truth choice, it receives a full accuracy reward of 1. If the letter is accurate but the response includes extra information, it is considered a partial match with a reward of 0.5. Incorrect answers, missing answers, or format violations result in an accuracy reward of 0.

The total reward *r*_*i*_ ∈ [0, 2] is computed as the sum of the format and accuracy rewards. This two-step reward structure ensures that the model first learns to produce interpretable, structured outputs before optimizing for clinical correctness.

### 2.1 Prompt Design

#### 2.1.1 MedVLM-R1 Prompt

MedVLM-R1 uses a general-purpose prompt format aimed at guiding the model through structured reasoning. The original prompt is as follows:

##### System Prompt

“A conversation between User and Assistant. The user asks a question, and the Assistant solves it. The Assistant first thinks about the reasoning process in the mind and then provides the user with the answer. The reasoning process and answer are enclosed within `<think>`…`</think>` and `<answer>`…`</answer>` tags, respectively.”

##### Question Template

“`{Question}` Your task:

1. Think through the question step by step, and enclose your reasoning process in `<think>`…`</think>` tags.
2. Then provide the correct single-letter choice (A, B, C, D) inside `<answer>`…`</answer>` tags.
3. No extra information or text outside of these tags.”

While effective in general settings, this format lacks clinical context and radiology-specific guidance. To address this limitation, we propose a revised prompt design that introduces structured medical reasoning, modality awareness, and anatomical precision. Our updated prompt is as follows:

#### 2.1.2 Proposed prompt

##### System Prompt

A conversation between a Radiology VQA Assistant and a User. The User provides a medical image-related question based on modalities like MRI, CT, or X-ray. The Assistant is a clinically informed expert model trained to analyze radiological images, identify visual features, reason step-by-step, and generate structured diagnostic responses. The Assistant always first performs detailed clinical reasoning, including image interpretation, anatomical localization, and potential differentials. This reasoning is enclosed in `<think>`…`</think>` tags. Then, the Assistant provides a final answer (A, B, C, D) enclosed in `<answer>`…`</answer>` tags. Do not generate any other text outside these tags.

##### Question Template

`{Question}` Your task:

1. Carefully analyze the radiological image and extract key visual features relevant to the question.
2. Perform clinical reasoning step-by-step, considering anatomy, pathology, and modality-specific characteristics.
3. Enclose your reasoning process inside `<think>`…`</think>` tags.
4. Then, provide the most appropriate single-letter answer (A, B, C, D) based on your reasoning, enclosed in `<answer>`…`</answer>` tags.
5. Do not include any extra explanation or information outside these tags.

This revised prompt structure introduces radiology-specific context, encourages step-by-step clinical reasoning, and constrains output formatting for improved evaluation and interpretability, as shown in Figure 1.

**Figure 1.**
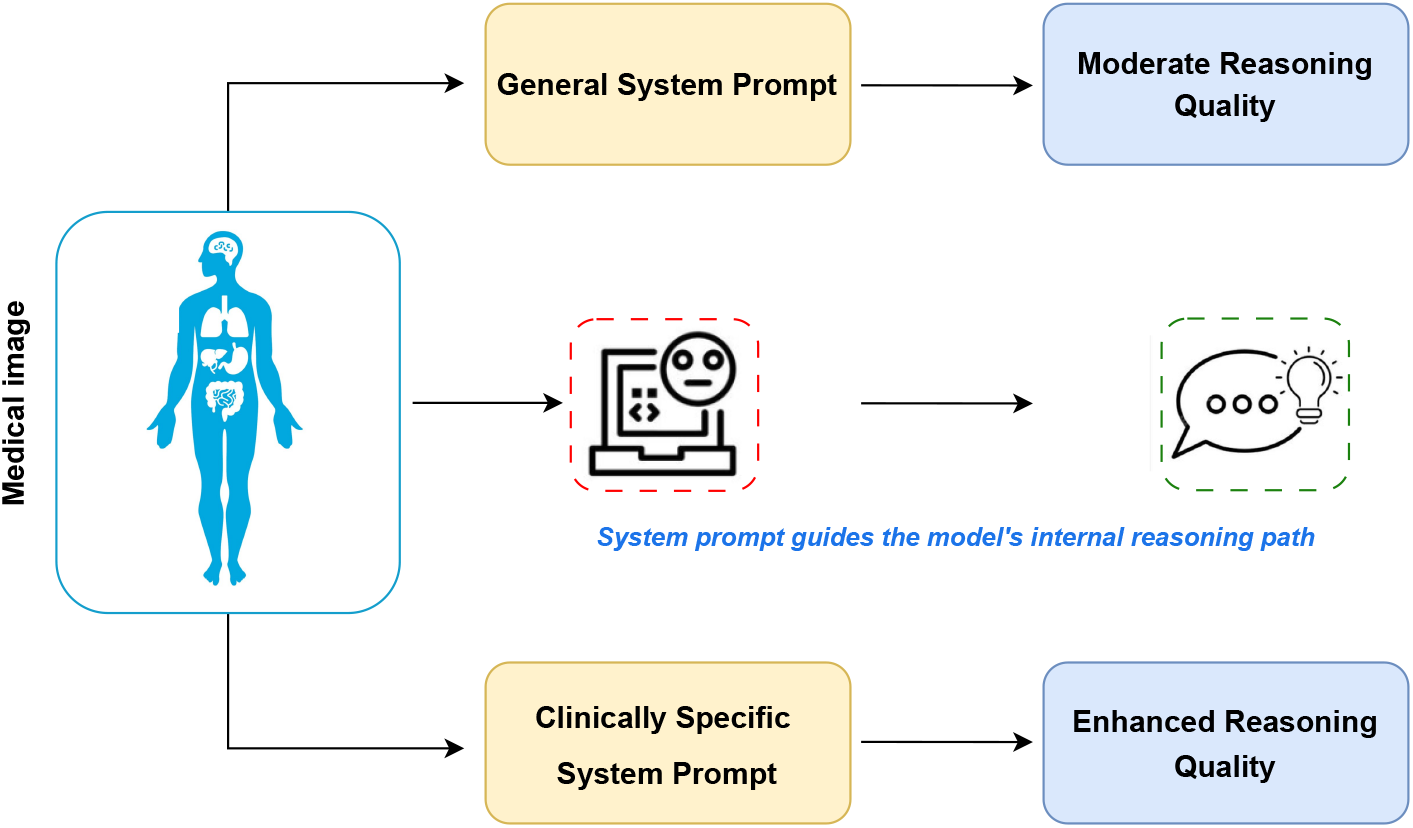
Block diagram of prompting and reasoning.

Figure 2 demonstrates the qualitative impact of our proposed clinically informed prompt design compared to the original MedVLM-R1 prompt. While both prompts yield the correct answer (B – Chest region), the explanation generated under the proposed prompt is noticeably more detailed and clinically grounded. The MedVLM-R1 prompt produces a generic anatomical reference, whereas our revised prompt encourages modality-specific interpretation (e.g., “chest CT scan”) and inclusion of relevant radiological features such as “lung nodules”. This example illustrates how structured, domain-specific instructions enhance the model’s diagnostic reasoning and reduce generic or underspecified outputs.

**Figure 2.**
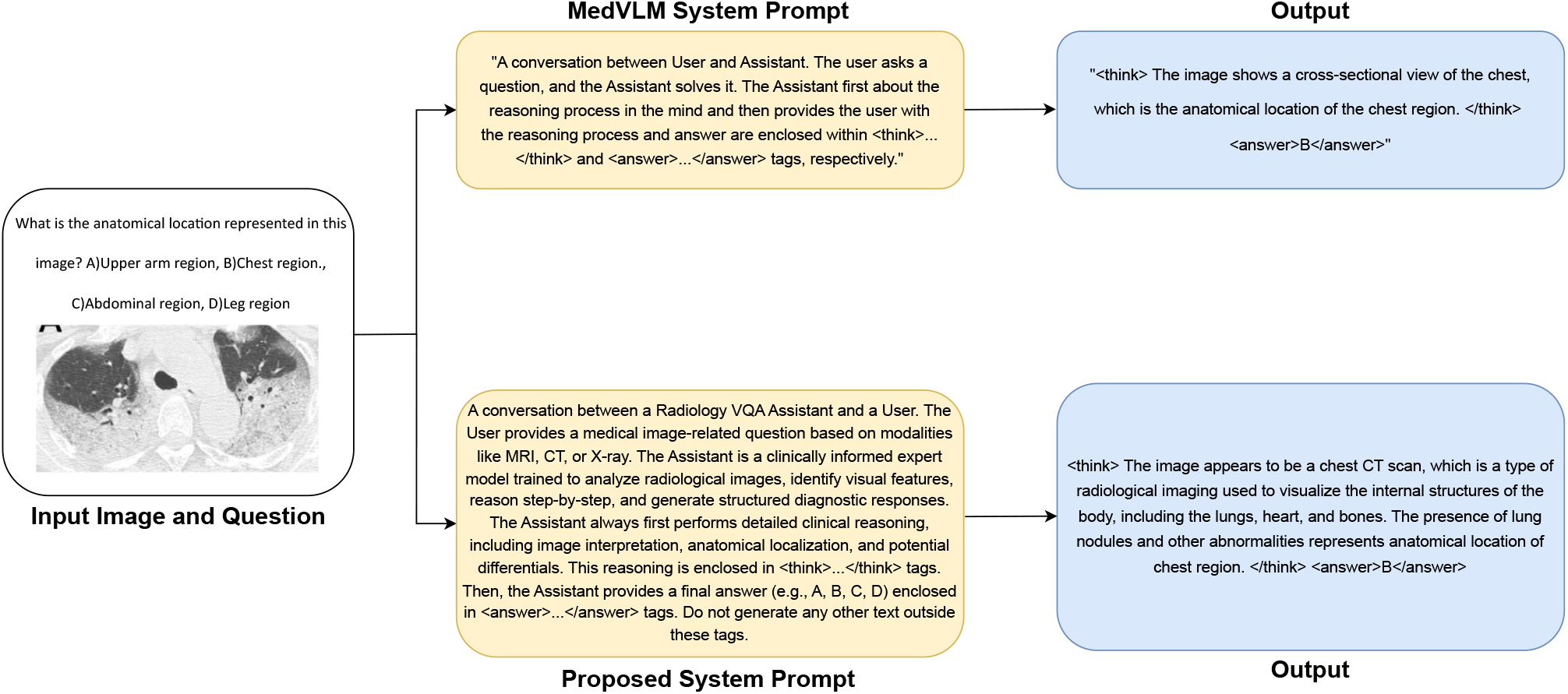
Example of improved reasoning with clinically informed prompt.

## 3 Experiment

### 3.1 Dataset Preparation

We utilize the HuatuoGPT-Vision dataset [28], a unified benchmark combining over 17,000 multiple-choice VQA samples from sources such as PMC-VQA [29], SLAKE [30], VQA-RAD [19], OmniMedVQA [31], and PathVQA [20]. These samples span diverse clinical tasks and imaging modalities, each paired with diagnostic images and two to six answer choices to reflect real-world medical decision-making.

In this study, we focus on three key modalities: MRI, CT, and X-ray. Training is conducted exclusively on 600 MRI image-question pairs, leveraging MRI’s structural complexity and rich diagnostic detail. To ensure fair comparison, our data split follows the same configuration as MedVLM-R1 in both size and task format. For evaluation, we use 300 MRI samples as in-domain tests and 300 samples each from CT and X-ray for out-of-domain (OOD) evaluation, as shown in Table 1. This consistent setup enables a controlled analysis of the model’s reasoning ability and its generalization across unseen modalities.

**Table 1.** Dataset composition for training and evaluation.

| Modality | Purpose | Samples | Type |
| --- | --- | --- | --- |
| MRI | Training | 600 | In-domain |
| MRI | Evaluation | 300 | In-domain |
| CT | Evaluation | 300 | Out-of-domain |
| X-ray | Evaluation | 300 | Out-of-domain |

### 3.2 Implementation

We fine-tuned MedVLM-R1, a vision-language model based on the Qwen2-VL-2B backbone [25], which was originally pretrained on a combination of curated web data, open datasets, and synthetic image-text pairs. While we preserve the original model architecture, reward structure, and training algorithm, we introduce a new radiology-specific system prompt and question template to better guide clinical reasoning and output structure. This modification enables us to evaluate the impact of domain-aware prompting while keeping all other training conditions identical.

Fine-tuning was conducted on two NVIDIA A100 80GB PCIe GPUs for 300 steps with a per-device batch size of 1. We used 6 candidate generations per input (*G* = 6), a maximum prompt length of 512 tokens, and a maximum response length of 512 tokens. All other training parameters, including learning rate, GRPO configuration, and reward functions, were kept identical to those used in the original MedVLM-R1 setup to ensure a controlled comparison.

## 4 Results and Discussion

We evaluate our prompt-augmented version of MedVLM-R1 on the HuatuoGPT-Vision dataset [28] across three different imaging modalities: MRI (in-domain), and CT and X-ray (out-of-domain). Our model was trained only on MRI samples, allowing us to assess its generalization capability to unseen modalities.

Table 2 shows that our prompt-tuned model achieves an accuracy of 96.00% on MRI, outperforming the MedVLM-R1 baseline by a small but consistent margin. More notably, we observe larger improvements on CT (+2.67%) and X-ray (+6.2%) modalities, despite no exposure to these during training. These results suggest that structured clinical reasoning prompts enhance the model’s generalization performance.

**Table 2.** Accuracy comparison between baseline MedVLM-R1 and our prompt-tuned model across imaging modalities.

| Model | MRI (In-Domain) | CT (OOD) | X-ray (OOD) |
| --- | --- | --- | --- |
| MedVLM-R1 | 95.33% | 70.00% | 69.00% |
| **Ours (Prompt-tuned)** | **96.00%** | **72.67%** | **75.20%** |

Qualitative analysis further reveals that the revised system prompt encourages more complete and medically grounded reasoning traces, reducing instances of hallucinated or underspecified explanations. The `<think>`…`</think>` segments in our model’s outputs consistently include modality-specific anatomical references, diagnostic pathways, and relevant differential considerations. For instance, in Figure 4, panel (a) illustrates the model identifying a meniscal abnormality in a knee MRI, with a detailed anatomical interpretation of the patella and surrounding tissues. In panel (b), the model correctly associates a CT image of the abdomen with liver disease and supports it with contextual knowledge about cross-sectional imaging and liver pathology. Similarly, panels (c) and (d) show that the model can accurately determine imaging modality (CT) and anatomical region (chest X-ray), while reasoning in a clinically consistent manner.

In contrast, the baseline MedVLM-R1, while fluent, often defaults to generic or template-like responses that fail to reflect the complexity of radiological interpretation, especially in OOD settings. This confirms that prompt design plays a crucial role in aligning model behavior with expert reasoning patterns, even without altering the model architecture or training dataset.

Figure 3 illustrates the training loss curve of the model over 300 optimization steps. The curve shows an overall decreasing trend, indicating progressive learning and convergence. While the majority of training steps yield low and stable loss values, several prominent spikes are observed, most notably around steps 85, 110, and 190. These transient increases in loss likely result from the inherent variance in reinforcement learning with group-based sampling, where challenging or diverse candidate responses can momentarily destabilize optimization. Despite these fluctuations, the model consistently recovers, with the final loss settling at a low value. These observations suggest that the fine-tuning process was effective and that the model successfully adapted to the training objective with improved alignment and output quality.

**Figure 3.**
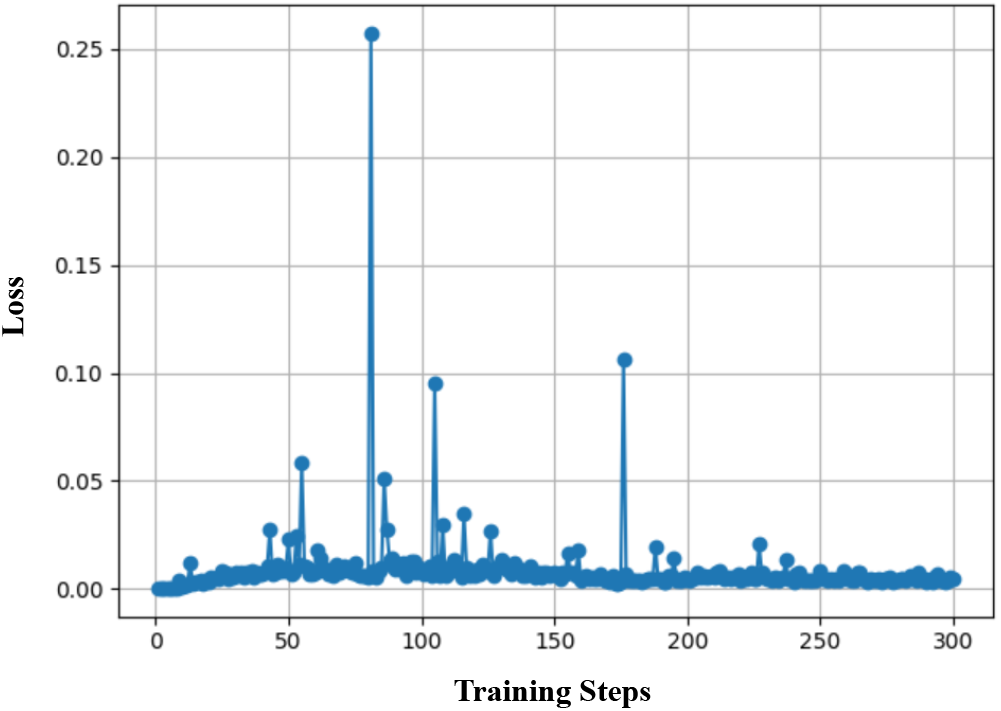
Training loss curve.

**Figure 4.**
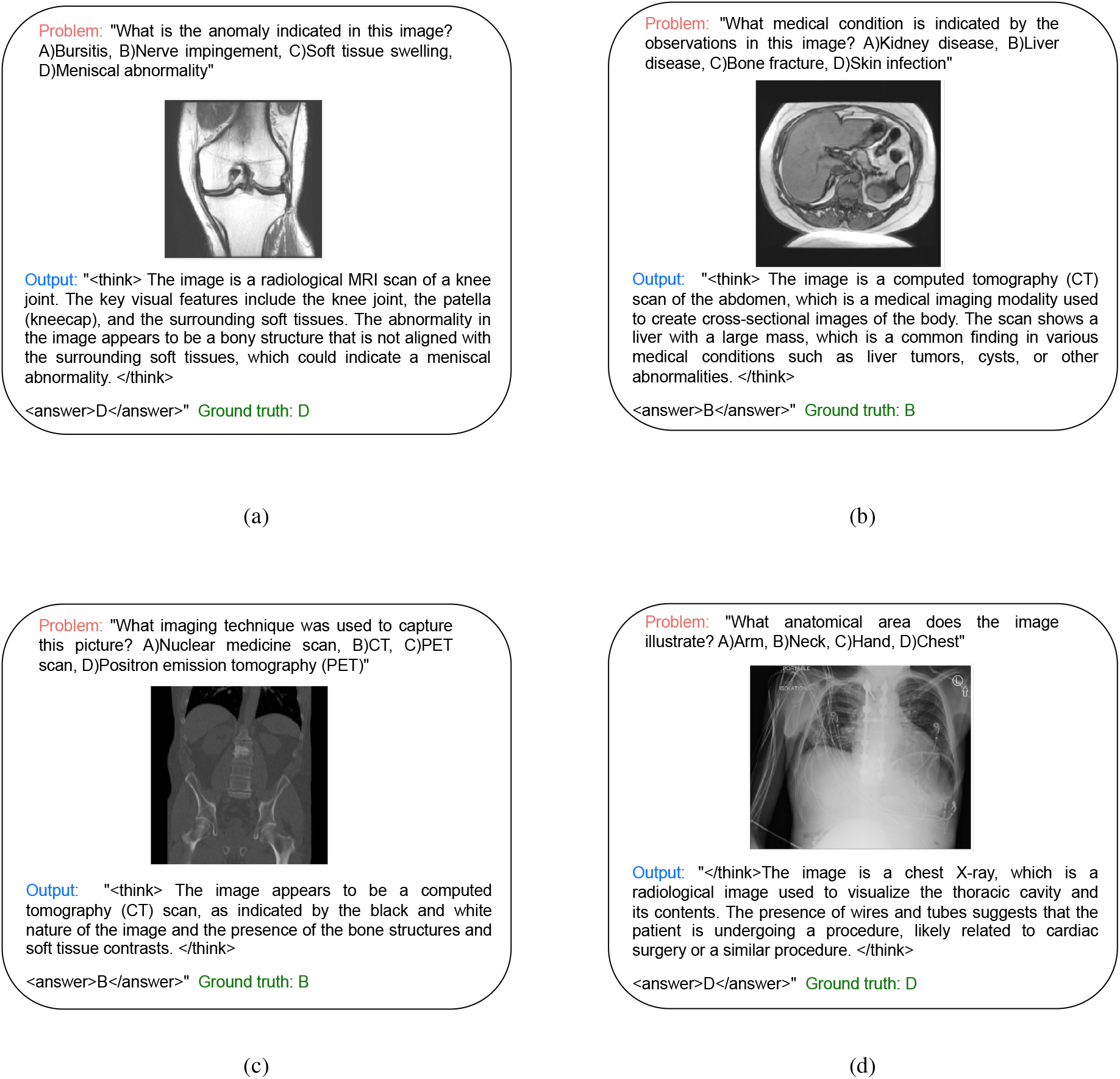
Medical VQA outputs of the proposed model across different modalities.

### 4.1 Limitations and Future Work

Although this study demonstrates the impact of prompt engineering in strengthening the diagnostic reasoning abilities of medical Vision-Language Models, several limitations remain. These reflect the scope of our current experiments and suggest key areas for future work. The model was fine-tuned only on MRI-based visual question answering samples. While this allowed a focused study on prompt design and enabled generalization tests on out-of-domain modalities like CT and X-ray, the lack of modality diversity during training limits the model’s ability to adapt to other imaging types. Including multi-modal training data could improve robustness and reduce bias toward MRI-specific features. The prompts used in this work were hand-crafted and static. Despite clear gains in interpretability and reasoning quality, such fixed templates may limit flexibility across varied medical tasks or user needs.

Future work could explore adaptive prompt strategies, such as instruction tuning, soft prompt learning, or reinforcement learning-based prompt generation, to support real-time adaptability. Our evaluation focused on multiple-choice VQA accuracy, which, while useful, does not fully capture clinical reasoning quality. Broader evaluation metrics, including hallucination detection, reasoning plausibility, and expert-rated interpretability, would provide a more complete performance picture. Lastly, the underlying model was pretrained on general-domain data. Further pretraining or adaptation using biomedical and radiology-specific corpora could improve alignment with clinical language and visual features, reducing errors from irrelevant associations.

In summary, this work highlights the promise of prompt-based alignment for improving structure and domain specificity in medical VLMs. Future efforts should address multi-modal training, adaptive prompting, comprehensive evaluation, and clinical pretraining to build transparent, reliable AI systems for healthcare. In future work, we will consider these limitations to further improve the model and develop more effective AI systems for medical applications.

## 5 Conclusion

In this study, we show that clinically informed prompts can significantly improve the reasoning capability and diagnostic accuracy of medical Vision-Language Models. Building upon the MedVLM-R1 framework, we introduced a radiology-specific prompt structure designed to encourage structured, step-by-step clinical reasoning. Without modifying the model architecture or optimization process, this prompt-tuned approach achieved improved performance across in-domain and out-of-domain tasks, notably surpassing the baseline model in both accuracy and interpretability.

Our findings reinforce the importance of input design in medical AI systems, particularly in high-stakes settings where hallucinations, under-explained answers, and lack of clinical grounding can limit model reliability. By embedding domain expertise directly into the prompt structure, we effectively aligned the model’s output format with expert expectations while preserving transparency and consistency in reasoning traces. yellow While the results are promising, this work also highlights areas for future exploration, including multi-modality training, dynamic prompt generation, and richer interpretability frameworks. As medical VLMs continue to evolve, such prompt-aware strategies will be essential for integrating general language models into robust and secure clinical decision support frameworks.

## Data Availability

All data used in this study are publicly available. The medical imaging data (MRI, CT, and X-ray) were obtained from the HuatuoGPT-Vision dataset, available at:
https://huggingface.co/FreedomIntelligence/HuatuoGPT-Vision-7B
No new data were generated in this study.

## 6 Acknowledgements

This work was supported by the research program of Korea Institute of Science and Technology Information (KISTI), the Korea Bio Data Station (K-BDS) with computing resources including technical support and the Institute of Information & Communications Technology Planning & Evaluation (IITP, under grant number RS-2024-00397359).

